# Inhibition of the Complement Pathway Induces Cellular Proliferation and Migration in Pancreatic Ductal Adenocarcinoma

**DOI:** 10.1101/2023.08.08.23293417

**Authors:** Zanele Nsingwane, Previn Naicker, Jones Omoshoro-Jones, John Devar, Martin Smith, Geoffrey Candy, Tanya Nadine Augustine, Ekene Emmanuel Nweke

**Affiliations:** Department of Surgery, Faculty of Health Sciences, University of the Witwatersrand, Johannesburg 2193. South Africa; Council for Scientific and Industrial Research, Pretoria. South Africa; Hepatopancreatobiliary (HPB) Surgery, Chris Hani Baragwanath Academic Hospital, Johannesburg. South Africa; School of Anatomical Sciences, Faculty of Health Sciences, University of the Witwatersrand, Johannesburg 2193. South Africa; Department of Life and Consumer Sciences, College of Agriculture and Environmental Sciences, University of South Africa, Florida, Roodepoort, South Africa

**Keywords:** PDAC, Immune response, cell migration, complement pathway, inflammation

## Abstract

Pancreatic ductal adenocarcinoma (PDAC) is a lethal cancer with a growing incidence and mortality despite novel therapeutic strategies. The complement signalling pathway may play diverse roles in PDAC by eliciting an immune response, inducing inflammatory responses, and may elevate pathways linked to chemoresistance. However, their role in the progression of PDAC is not fully understood. In this study, 30 tissues and 34 plasma samples were obtained from a cohort of PDAC patients including controls. Targeted pathway-specific PCR analysis was conducted to determine the gene expression profiles of immune-response-related genes. The circulating levels of complement proteins C3 and C5 were further investigated. Pharmacological inhibition of the complement pathway in MIA PaCa-2 pancreatic cancer cell lines was performed and the effect on cells was assessed by cell proliferation, cell migration, and cell cycle assays. Finally, SWATH-mass spectrometry was performed to identify potential molecular mechanisms during inhibition. The results identified C3 and C5 to be overly expressed in early PDAC compared to later stages. Pharmacological inhibition of the complement pathway led to increased cell growth, proliferation and migration *in vitro*. Proteomic analysis implicated several proteins such as the mitochondrial and histone proteins, that could play a role in inducing this phenotype. This study helps to further delineate the role of the complement pathway in PDAC progression suggesting a context-dependent function.

## Introduction

Pancreatic ductal adenocarcinoma (PDAC) is a lethal cancer with a growing incidence and mortality despite current therapeutic strategies (1). It is widely regarded as non-immunogenic due to its immunosuppressive tumour microenvironment (2). Recent advances in cancer immunotherapy have been successfully achieved in numerous cancers, such as melanoma and some lymphomas, but have not yielded improved results in PDAC. Pancreatic cancer is known to be associated with inflammation, and the immune response plays a crucial role in the development and progression of the disease. Inflammatory signals can promote the growth and spread of cancer cells by creating an environment that supports tumour progression (3). Understanding the interplay between inflammation and immune response in pancreatic cancer is essential for developing effective therapeutic strategies.

The complement signalling pathway, which involves the activation of specific complement proteins, plays a vital role in immune surveillance and is responsible for facilitating the elimination of invading pathogens and ‘non-self’ antigens (4). Once activated, these complement proteins can trigger a cascade of events that ultimately lead to the destruction of foreign substances. Aberrations in this pathway are associated with chronic inflammation, cancer, and autoimmune diseases (5). This pathway can be activated via three main routes: classical, mannose-binding lectin and alternative pathways, with all having the complement C3 as a central protein. C3 functions as a master switch that regulates all downstream factors and proteins, including the complement C5, responsible for pathogen elimination via the membrane attack complex (MAC). Understanding the complement system’s role in cancer has evolved over the years, with complement proteins now implicated in several hallmarks of cancer (6,7). Although the complement pathway is involved in eliciting immune response, it may also play a tumour-promoting role (8). Some studies have suggested that activation of the complement pathway can result in increased inflammation, fostering a favourable environment for cancer cell growth and metastasis. In PDAC, increased complement protein levels have been demonstrated to elevate anaphylatoxins, C3a and C5a, which directly induce TNF-alpha and IL-1B (4). Furthermore, C3 and C5 are linked to chemoresistance by potential interactions with several pathways such as nuclear factor kappa B (NF-kB), phosphoinositide-3-kinase/protein kinase B (PI3K/AKT), C-mesenchymal-epithelial transition factor (C-MET) and the signal transducer and activator of transcription (STAT3) pathways (4,9). However, the mechanisms underpinning these interactions are not fully understood.

In this study, tumour and plasma samples from PDAC patients were profiled using immune-response pathway-specific PCR arrays, real-time PCR and ELISA. The levels of complement proteins C3 and C5 were confirmed to vary with tumour severity. A pancreatic cancer cell line model (MIA PaCa-2) was used to demonstrate the effects of the pharmacological inhibition of the complement pathway. Additionally, SWATH mass spectrometry was conducted to identify proteins dysregulated following inhibition of the complement pathway.

## Methods

### Patient selection and recruitment

Patients were recruited from the Hepatopancreatobiliary (HPB) unit of Chris Hani Baragwanath Academic Hospital (CHBAH) in Johannesburg between January 2017 and March 2020. Recruited patients were self-identified black Africans and were between the ages of 18 and 80. Patients presenting with acute inflammation, sepsis, organ failure and those undergoing therapy were excluded from the study and were not eligible to participate. All PDAC patients were clinically and histopathologically diagnosed with pancreatic ductal adenocarcinoma (PDAC) following a screening of the abdomen using a contrast-enhanced triple-phased CT scan. Histopathological diagnosis was by either endoscopic or surgical means. Endoscopic pathological diagnosis was by brush cytopathology of structures at endoscopic retrograde-cholangiopancreatophy (ERCP), or biopsy of pancreas mass by endoscopic ultrasonography (EUS).

Tissues: Biopsy samples (a tumour and corresponding normal tissue) from fifteen (15) patients (10) scheduled for a Whipple procedure (surgical resection of cancerous tumours located on the head of the pancreas), were immediately immersed in RNA *later*™ stabilisation solution (InvitrogenÔ, Massachusetts, United States).

Plasma: A separate cohort of twenty-five PDAC patients (Thirteen resectable, eight locally advanced and four metastatic) categorised according to the National Comprehensive Cancer Network guidelines (NCCN) (11) and six chronic pancreatitis patients were recruited. Individuals (three) who self-reported being in good health and not taking any medication were recruited as healthy control participants (Table S1). Whole blood (10ml) was collected into EDTA vacutainer tubes (BD Biosciences, Franklin Lakes, NJ, USA) by venepuncture and processed within two hours of collection. The blood was separated by gravity separation and then centrifuged at 3000xg at 4°C for 30 minutes, and the isolated plasma was aliquoted (500 µl) into Eppendorf tubes. All samples were stored in a −80° C freezer until required, and analysis was conducted within six months.

### Total RNA extraction from tissues and cells

Total RNA was extracted using the TRIzol method (12). Tissues (15 – 25mg) were shredded into small pieces using sterilised surgical blades and transferred into 15ml Falcon tubes containing 700 μl of TRIzol reagent (Sigma Aldrich, St. Louis, Missouri, United States) and then homogenised using a TissueRuptor (Qiagen, Hilden, Germany). For the MIA PaCa-2 cells, a cell pellet was resuspended in 700 μl of TRIzol homogenous mixture, transferred into 1.5ml Eppendorf tubes and allowed to stand for five minutes at room temperature to ensure nucleoprotein complexes were fully dissociated. Chloroform (150μl) (Merck, Darmstadt, Germany) was added and shaken vigorously for 15 seconds until the mixture turned milky and left to stand for eight minutes before being centrifuged at 12 000xg for 10 minutes at 4° C. The aqueous phase was transferred into sterile Eppendorf tubes, and 350 μl of isopropanol (Sigma Aldrich St. Louis, Missouri, United States) was added to the sample, mixed by resuspension and allowed to stand for seven minutes. The mixture was centrifuged at 12 000xg for eight minutes at 4° C. The supernatant was discarded, and the pellet was washed with 700μl of 75% cold ethanol (Sigma Aldrich St. Louis, Missouri, United States). The sample was vortexed for five seconds and centrifuged at 7500xg for five minutes at 4° C. The supernatant was discarded, and the pellet was air-dried for 10 minutes and placed into a dry bath (55°C) for further drying. A total of 30μl nuclease-free water (Sigma Aldrich St. Louis, Missouri, United States) was added to the pellet, mixed repeatedly with a pipette, placed in the heating block for five minutes at 55^°^C, and immediately placed on ice. Total RNA concentration and quality were assessed using the Nanodrop® 1000 (Thermo Fisher, Waltham, Massachusetts, United States). The A260/A280 and A260/230 of samples were all over 2.0.

### Pathway-focused PCR array analysis

The RT^2^ first strand kit (Qiagen) was used for cDNA synthesis as per the manufacturer’s instructions from two micrograms of total RNA. To perform a pathway-focussed gene expression analysis of immune response genes, the RT^2^ Profiler PCR Human Innate and Adaptive immune responses (PAHS-052ZA) panel were used. The PCR mix was prepared as per the manufacturer’s instructions and ran on the QuantStudio1 real-time thermocycler (Thermo Fisher Scientific). The Ct values were exported and fold changes were analysed using the QIAGEN RT^2^ Profiler PCR analysis software tool (https://www.qiagen.com/us/product-categories/instruments-and-automation/analytics-software accessed 26 July 2023).

### Verification of gene targets using real-time PCR

Verification of gene targets was achieved by performing cDNA synthesis followed by real-time PCR according to the MIQE guidelines (13). The Invitrogen^TM^ SuperScript^TM^_VILO^TM^ cDNA synthesis kit (ThermoFisher Scientific) was used followed by real-time PCR. The method involved adding five microlitres of TaqMan^®^ Gene Expression Master Mix (2X), 0.5µl TaqMan^®^ Gene Expression Assay 20X) for target gene and reference gene, three microlitres of cDNA template, Human Male Raji cDNA template (controls), and one microlitre nuclease-free water to a PCR plate, briefly vortexed, centrifuged and run according to the manufacturer’s instructions (Thermo Fisher Scientific). The targets were *C3* and *C5* with *MRPL-19* as a reference gene, Fold changes were determined using the 2^−ΔΔCT^ method (14).

### Enzyme-linked immunosorbent assay (ELISA) analysis

The human complement C3 and C5 ELISA kits (Abcam, Cambridge, United Kingdom) were used to quantitatively measure plasma C3 and C5 across PDAC patients in different stages (resectable, locally advanced and metastatic), chronic pancreatitis patients and healthy individuals. Based on the manufacturer’s recommendation, plasma samples were diluted 1:800 into 1X Diluent M for C3 and 1:20 000 into 1X Diluent N for C5. 1X wash buffer, 1X Streptavidin-Peroxidase Conjugate, 1X biotinylated complement C3/C5, seven standards and a blank were prepared according to the manufacturer’s protocol.

Twenty-five microlitres and 50 ml of samples and standards were added to each well of a pre-coated and blocked 96-well microtiter plate for C3 and C5. For C3, 25µl of 1X biotinylated complement C3 antibody was added and incubated at RT for 2 hrs. For C5, 50µl of 1X biotinylated complement C5 antibody and incubated for 1 hr, Plates were washed as per manufacturers guidelines followed by the addition of 50µl 1X Streptavidin-Peroxidase Conjugate to all wells for 30 minutes at RT. Stop solution (50µl) was added to each well, and the colour change from blue to yellow and absorbance was measured at 450nm immediately using the Biochrom Anthos 2010 Microplate reader (Biochrom Ltd, Cambridge, United Kingdom).

### Cell culture and treatment

The pancreatic cancer cell line, MIA PaCa-2, representing a cancer model, was used in this study. MIA PaCa-2 cells were derived from a 65-year-old Caucasian male patient with pancreatic adenocarcinoma located at the head and tail of the pancreas (15). The MIA PaCa-2 were grown in Dulbecco’s Modified Eagle Medium (DMEM) (Sigma Aldrich) with 10% foetal bovine serum (FBS) (Sigma Aldrich) and 1% penicillin-streptomycin (Sigma Aldrich) and maintained in an incubator at 37 °C in 5% CO_2_ and 95% air. Compstatin (Tocris Bioscience, Bristol, United Kingdom) was used to inhibit the complement pathway pharmacologically, and dimethyl sulfoxide (DMSO) (Sigma Aldrich) as vehicle control. Viable cell counts were determined using the trypan blue exclusion assay, by adding 10µl cells to trypan blue (1:1 ratio) to the cell suspension and counting on a TC20^TM^ automated cell counter (Bio-Rad Laboratories, Hercules, California, USA)

### Cell proliferation assay using methoxynitrosulfophenyl-tetrazolium carboxanilide (XTT)

The cell proliferation kit II (XTT) (Sigma Aldrich) colourimetric assay was employed to quantify cell proliferation, viability and cytotoxicity in pancreatic cancer MIA PaCa-2 cells. Cells were cultured and viability determined as described above). Viable MIA PaCa-2 cells (2.5×10^3^, 5×10^4^ and 1×10^5^ cells/well)were seeded in different 96-well plates (ThermoFisher Scientific) in 200µl complete medium and incubated at 37°C, 5% CO_2_ for 24 hours in triplicate. Once cells reached 60-70% confluence, they were treated with a range (0–100µM) of complement inhibitor (Compstatin). Controls included a negative control (normal DMEM + 10% FBS + 1% pen-strep and cells), the vehicle control (0.1% DMSO in complete medium), cells treated with gemcitabine as a positive control and just medium with no cells as a blank) for 24 and 48 hours. Absorbance was measured on a spectrophotometer (MULTISKAN Sky, ThermoFisher Scientific) at 450 nm with a reference wavelength of 690 nm. Percentage survival for each sample was calculated as 100LJ×LJ[(OD450 of sampleLJ−LJOD450 of negative control)/(OD450 of positive controlLJ−LJOD450 of negative control)].

### Cell cycle analysis

Measurements of the percentage of cells in the G0/G1, S and G2/M phases of the cell cycle were based on analysing DNA content using the single parameter-based propidium iodide staining technique (BD Biosciences, New Jersey, USA). MIA Paca-2 cells were counted and 2 x 10^6^ viable cells were seeded in duplicate and left to grow for 24 hours. Cells were treated with 6µM Compstatin to inhibit the complement pathway and with 0.1% DMSO as vehicle control. Following trypsinisation, cells were centrifuged at 250xg for five minutes, fixed in cold 70% ethanol and stored at −20°C for at least 24 hours. For permeabilization and removal of RNA, 500µl of 2mg/ml RNAse A in 0.1% Triton X-100 in PBS (BD Biosciences) and 400µl of 500µg/ml propidium iodide dye solution in PBS, were resuspended with 100 µl of the sample. The suspension was incubated at room temperature for 30 minutes, and fluorescence was measured using the BD LSR Fortessa™ Analyser and FACS Diva software (BD Biosciences). The cell cycle phase was estimated using FlowJo™ v.10 software (BD Biosciences).

### Cell migration (scratch assay)

Viable MIA PaCa-2 cells (1 x 10^5^), counted as previously mentioned, were seeded in a 24-well plate (ThermoFisher Scientific) by adding 1 x 10^5^ µl cell and media solution to each well in triplicate. Plates were incubated at 37°C, 5% CO_2,_ for 24 hours. Once cells reached 80-100 % confluence, the cell monolayer was imaged using the Olympus iX51 phase contrast microscope (Wirsam Scientific & Precision Equipment Ltd, Johannesburg, South Africa), followed by washing cells twice with 0.1M PBS. A scratch wound was made across the wells on the second wash using a sterile 10µl pipette tip. The cells were then washed three times with PBS. To redirect cellular focus from proliferation to migration while maintaining viability, cells were exposed to a low serum medium supplemented with 1% Fetal Bovine Serum (FBS) instead of the standard 10% FBS (16) to reduce nutrient levels, allowing the cells to prioritize migration over proliferation, albeit with some residual proliferation activity. Cells were treated with 6µM Compstatin to inhibit complement activation; 0.1% DMSO was used as vehicle control. Images of the scratched area were captured using the Olympus iX51 phase contrast microscope with CellSens Software (Wirsam Scientific & Precision Equipment Ltd) at 0 h, 6 h 12 h and 24 hrs. The following calculation was used to measure the rate of cell migration (D_initial–_ D_final_) 100/ D_initial_, where D is distance.

### Proteomic analysis using SWATH-MS

MIA PaCa-2 cells were counted and 1 x 10^5^ viable cells were seeded in duplicate in a 24-well plate and left to grow for 24 hours before being treated with 6 µM Compstatin to inhibit the complement pathway by targeting C3. A cell pellet was prepared by washing adherent cells with PBS and adding trypsin-EDTA to detach the cells. Cells were by centrifuging at 250xg for five minutes. The pellet was washed at least four times with PBS and a dry pellet was stored at −80° C until processing. Treated and untreated cell pellets were resuspended in 50mM Tris-HCl pH 8.0, containing 2% SDS. The protein concentration was measured using the Pierce Bicinchoninic assay (ThermoFisher Scientific), per the manufacturer’s instruction. Protein samples (10μg per sample) were reduced with 5mM tris (2-carboxyethyl) phosphine and alkylated with 10mM 2-chloroacetamide at room temperature for 20 minutes. Samples were purified of detergents and salts using MagReSyn™ HILIC beads (ReSyn Biosciences) as previously described (10,17). On-bead protein digestion was performed using a 1:20 and 1:100 protease: protein ratio for sequencing-grade trypsin and Lys-C, respectively. The resultant peptides were dried and stored at −80°C before LC-MS analysis.

Approximately 1µg of peptides per sample was analysed using a Dionex Ultimate 3000 RSLC system coupled to a Sciex 5600 TripleTOF mass spectrometer. Injected peptides were inline de-salted using an Acclaim PepMap C18 trap column (75μM × 2cm: 2 min at 5μl.min^-1^ using 2% ACN/0.2% FA). Trapped peptides were gradient eluted and separated on a Waters Acquity CSH C18 NanoEase column (75μM × 25cm, 1.7µm particle size) at a flow rate of 0.3 µl.min^-1^ with a gradient of 6-40% B over 90 minutes (A: 0.1% FA; B: 80% ACN/0.1% FA). For sequential window acquisition of all theoretical mass spectra (SWATH), precursor scans were acquired from 400-1100*m/z* with 50 msec accumulation time and fragment ions were acquired from 200-1800*m/z* for 48 variable-width precursor windows with 0.5 Da overlap between windows and 20 milliseconds accumulation time per window.

SWATH data was processed using Spectronaut v17 software (Biognosys). The default directDIA identification and quantification settings were used for data processing. Carbamidomethylation was added as a fixed modification, and N-terminal acetylation and methionine oxidation were added as variable modifications. Swiss-Prot Human sequences (downloaded on 03 March 2023 from www.uniprot.org) and common contaminating proteins were used as the search database. A q-value ≤ 0.01 cut-off was applied at the precursor and protein levels. Quantification was performed at the MS1 and MS2 levels. Label-free cross-run normalisation was employed using a global normalisation strategy. Candidate dysregulated proteins were filtered at a q-value ≤ 0.01 and absolute Log_2_ fold change (FC) ≥ 0.58.

### Statistical and Bioinformatics analyses

The Kruskal-Wallis and median tests were used to compare differences between groups due to a small sample size, and the Mann-Whitney U test was performed for posthoc analysis. Spearman correlation analyses were also conducted and the differences were regarded as significant at p<0.05 in all tests. Functional enrichment analyses were performed on all the dysregulated proteins. Interaction network analysis and visualisation were performed using Cytoscape v3.8.2 (18). The StringApp v2.0.1 (19) and Reactome v8.0.5 (20) plugins on Cytoscape were used to determine the molecular functions, cell compartments and pathways enriched by the dysregulated proteins. The proteins were queried with filters including *homo sapiens* as the reference species, a confidence cut-off score of 0.4 and zero additional interactors. Single non-interacting proteins were excluded from the interaction network.

## Results

### Dysregulated immune response genes in pancreatic tumours

The gene expression profile of immune response genes was investigated in pancreatic tumours in comparison to corresponding normal tissues. A total of 13 and 62 genes upregulated and downregulated, respectively (Figure 1a). *C3* was the most upregulated gene (Fold change = 562.22) and chemokine (C-C motif) receptor 4 (*CCR4*) was the most downregulated gene (Fold change = −83.19). Furthermore, among the top five upregulated targets, three were major histocompatibility complex molecules, and their expression levels were 20-100 times lower compared to the *C3*.

**Figure 1:**
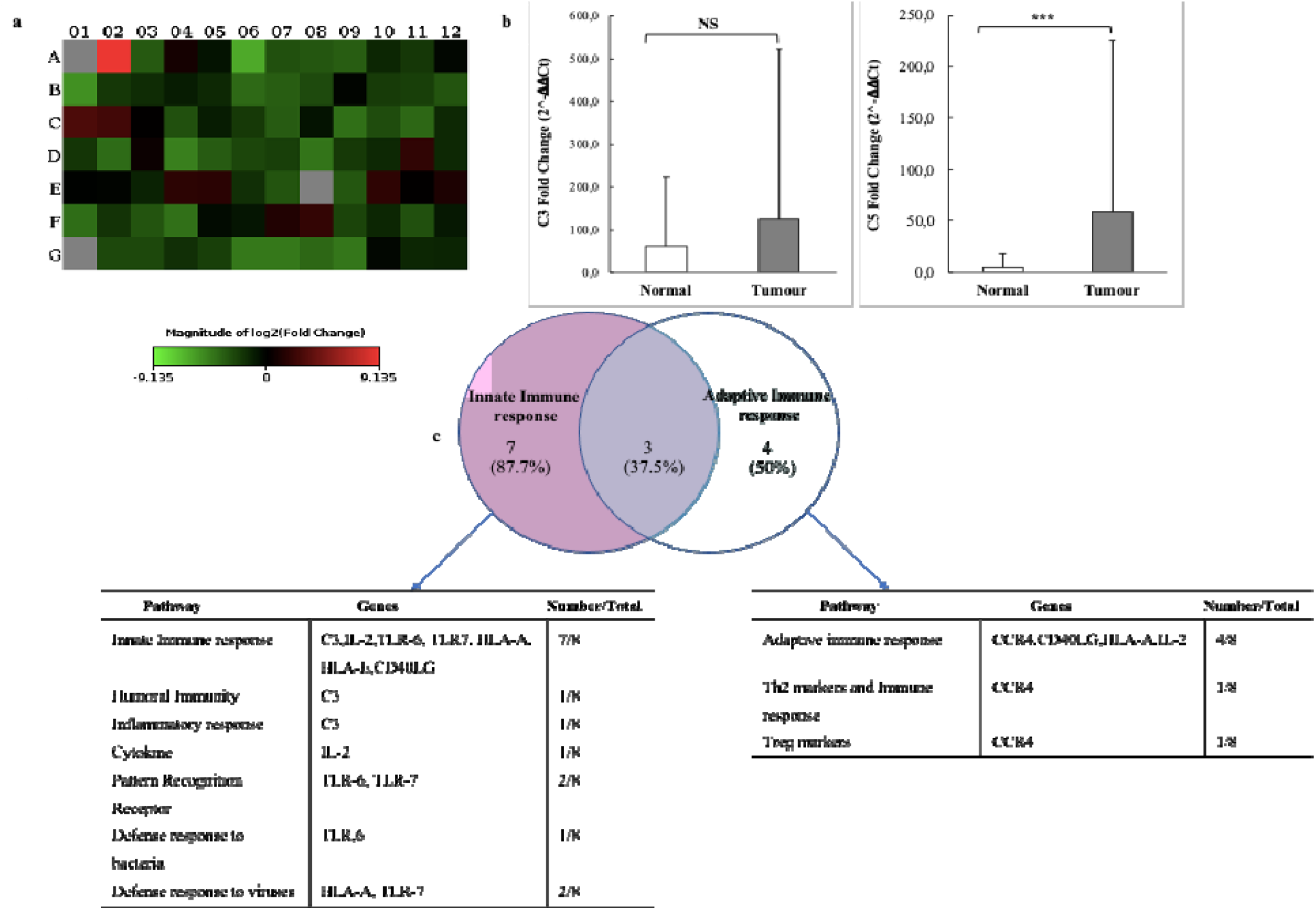
Gene expression profiles of immune response genes in PDAC tumours. (a) Heatmap showing differentially expressed genes in a form of fold regulation. Red represents upregulated genes with positive values, and green represents downregulated targets. (b) Real-Time PCR verifying the gene expression profile of complement C3 and C5 in resectable PDAC tumours. C3 and C5 are upregulated in pancreatic tumours and downregulated in normal tissues. (NS - non significant, *** indicate - p<0.0001). (c) The Venn diagram shows that most genes function predominantly in the innate immune response but also have some role in the adaptive immune response.

Real-time PCR was employed to validate the expression of the complement *C3* in a larger sample group as samples were pooled previously for the RT2 profiler PCR array to minimise costs. Although the increase of the complement C3 in pancreatic tumours compared to normal pancreatic tissue was confirmed, it was not statistically significant (p=0.72).

We further evaluated the expression of another crucial downstream component of the complement pathway, *C5*, which was not included in the PCR array. A significant increase in the expression of complement *C5* in pancreatic tumours compared to normal pancreatic tissues was observed (p<0.0001) (Figure 1b).

Moreover, on examining the top five genes that were upregulated *(C3, B2M, HLA-A, RPLP0, HLA-E)* and downregulated *(CCR4, CD40LG, IL-2, TLR-7, TLR-6),* it was observed that these genes function primarily in the innate and adaptive immune responses but are also involved in part of the adaptive immune response based on their classification in the array panel (Figure 1c).

### Complement C3 and C5 are elevated in the plasma of early-stage PDAC patients

The levels of C3 and C5 were investigated in the plasma of a cohort of PDAC patients at different disease stages and compared to chronic pancreatitis patients and healthy individuals. There were no observed significant changes across the different tumour stages; however, a significant (p=0.0471) reduction in C3 levels was observed when patients with early-stage (resectable) cancer were compared to those with advanced-stage disease (locally advanced and metastatic) (Figure 2a). Of note, the expression of C3 was at its highest level in patients with chronic pancreatitis compared to other groups. The levels of C5 were increased in all patient groups, including cancer and chronic pancreatitis patients, compared to healthy individuals. Additionally, C5 was shown to be significantly elevated in patients with resectable PDAC compared to healthy individuals (p=0.0155) (Figure 2b). Although there was a reduction in its levels in late-stage disease compared to early-stage, this was not significant. Furthermore, additional statistical analysis demonstrated a positive correlation (r=0.45669) between the complement C5 and the clinical inflammatory marker C-reactive protein (CRP) (p = 0.0098) (Figure S1).

**Figure 2:**
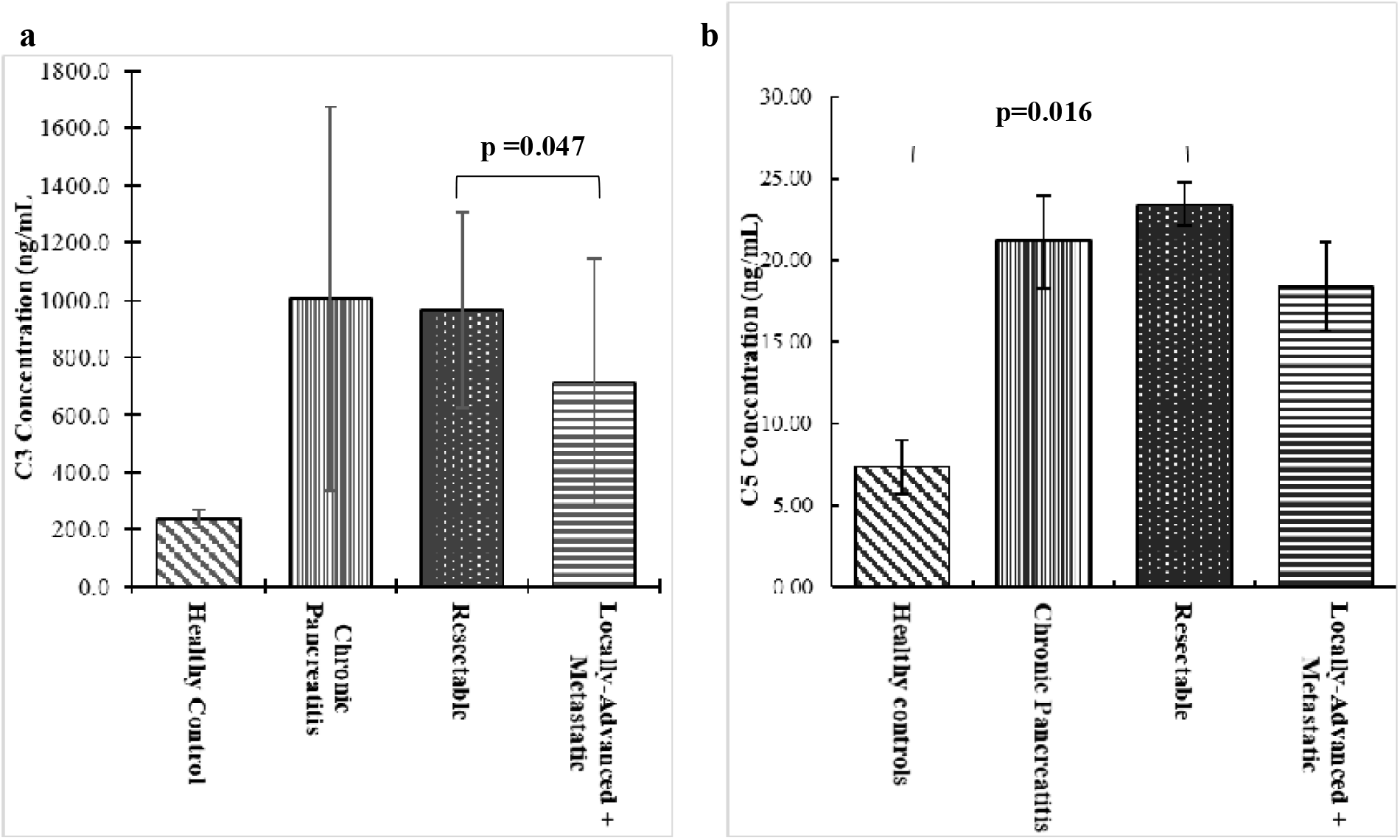
Protein expression levelsC3 and C5 in plasma of PDAC patients. (a-b) A decreasing trend is observed in all PDAC samples from early-stage to late-stage disease. Healthy individuals express low quantities of C3 and C5.

### Pharmacological inhibition of complement pathway induces cell proliferation, cell division and migration of MIA PaCa 2 cells

To investigate further the implications of reduced complement pathway activation, cells were treated with Compstatin and cell-based analyses were performed. First, the inhibition of the complement pathway was confirmed by a reduction of C3 and C5 levels (Figure S2) after 24 hours of treatment with 6 µM compstatin (Figure 3a).

**Figure 3:**
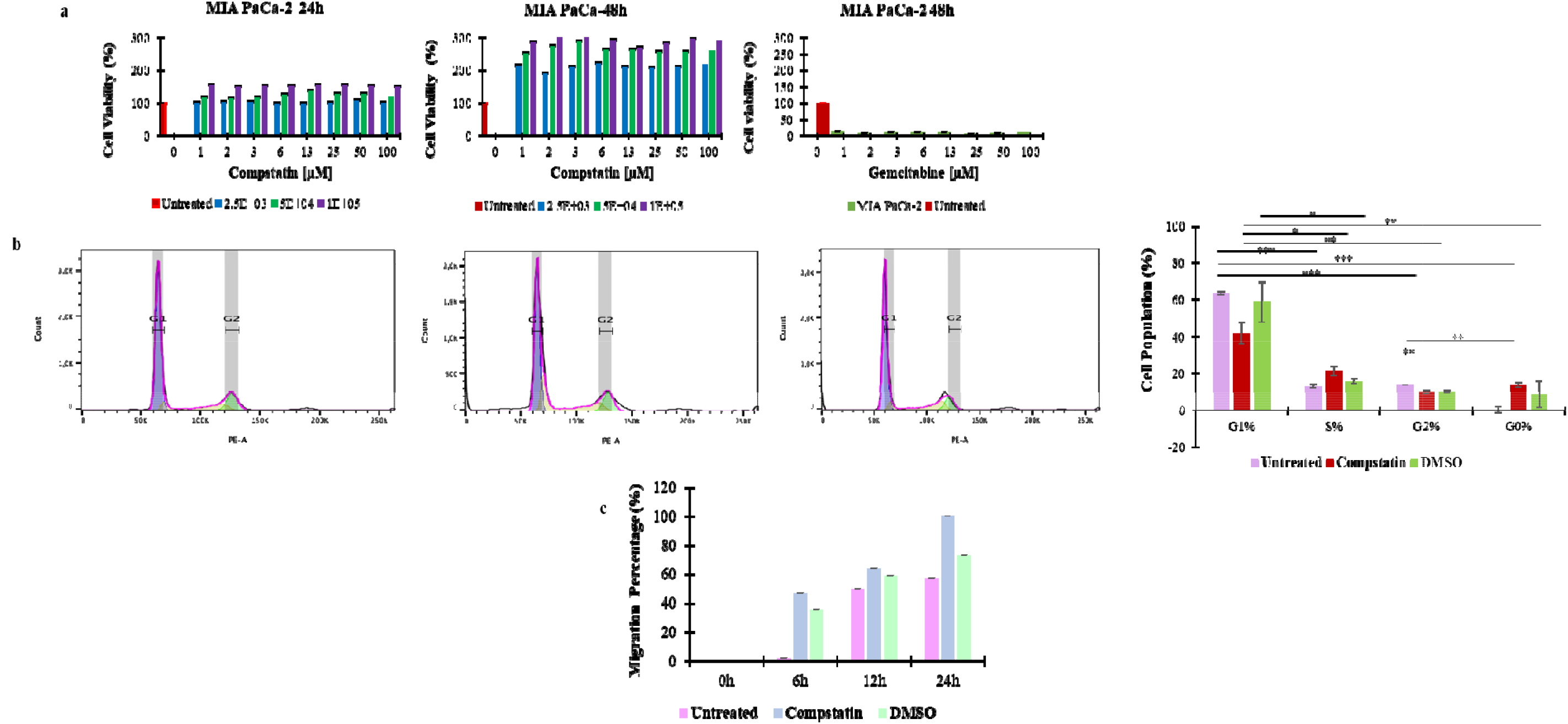
Enhanced Cell Proliferation and Migration Induced by Complement Inhibition in MIA PaCA-2 cells. (a) Inhibition of the complement pathway induced cell proliferation as evidenced by increased viability (>100%) in MIA PaCa-2 cells over 48hrs, whereas gemcitabine was toxic to MIA PaCa-2. (b Complement inhibition allowed cells to transition from G1 to S phase showing increased cell division. (c) Complement inhibition enhances cell migration, compared to both the untreated and DMSO vehicle controls

The XTT cell proliferation assay showed that complement C3 inhibition resulted in a significant increase (p=0.001) in the number of viable MIA PaCa-2 cells, thus indicative of heightened cell proliferation, compared to untreated cells (Figure 3a). In contrast, treatment with gemcitabine, used as a positive control, led to a reduction in cell viability in both normal and cancer cells.

Flow cytometry was used to investigate the effect of complement inhibition on the cell cycle of MIA PaCa-2 cells. The findings showed that untreated cells were significantly higher (p<0.0001) in G1 phase and lower G0 phase. However, after compstatin treatment, the cells exhibited a significant reduction in G1 phase and an increase in S phase (p=0.004) necessary for DNA replication and repair, and also increased in G0 phase, though the increase was not statistically significant (p=0.5055) (Figure 3b).

The scratch assay showed complement inhibition accelerates cell migration in MIA PaCa-2 cells (Figure 3c).

### Proteins dysregulated with inhibition of the complement pathway

To further delineate the potential molecular mechanisms linked with the inhibition of the complement pathway in PDAC, MIA PaCa-2 cells were exposed to compstatin for 24 hours. Following treatment, SWATH-MS was used to conduct a proteomic analysis and the results identified a total of 2814 protein groups. A comparison of the compstatin-treated cells with untreated cells showed that there were 53 upregulated proteins and 60 downregulated proteins (Table S2). Furthermore, the upregulated proteins included a variety of crucial proteins such as the Cysteine and Glycine Rich Protein 1 (CSRP), Mitochondrial Ribosomal Protein (MRPL4, MRPL47), Mitochondrial Ribosome Recycling Factor (MRRF), Glutamate Cysteine Ligase catalytic subunit (GCLC), Euchromatin Histone Methyltransferase II (EHMT2) and RNA Polymerase I and III subunit D (POLR1D). Moreover, the H2A Clustered Histone family of molecules (H2AJ, H2AC7, H2AC12, H2AC14, H2AC17, H2AC19 and H2AC20), were among the top five downregulated proteins (Figure 4). The two main pathways that were impacted were the upregulation of the MTF1-activated gene expression pathway and the downregulation of the RMTs methylate histone arginines pathway (Table 1).

**Figure 4:**
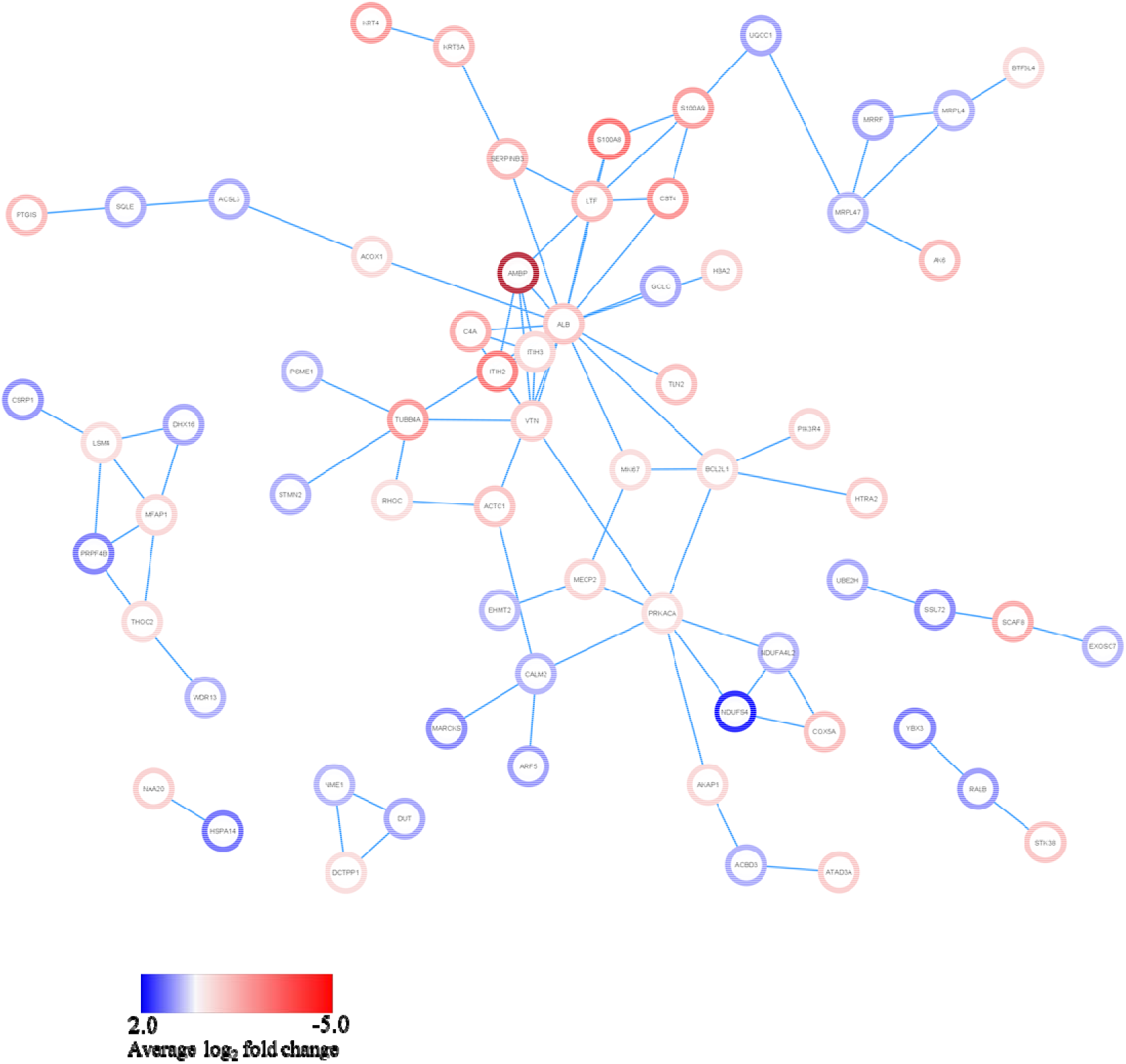
Network analysis showing the intricate interaction of dysregulated proteins. Blue and red represent upregulated and downregulated proteins, respectively in compstatin-treated vs untreated pancreatic cancer cells

**Table 1:** Top 10 up- and down-regulated pathways.

| Upregulated |  |  | Downregulated |  |  |
| --- | --- | --- | --- | --- | --- |
| Pathway name | p-value | Proteins enriched | Pathway name | p-value | Proteins enriched |
| MTF1 activates gene expression | 3.95e-04 | CSRP1 | RMTs methylate histone arginines | 7.96e-10 | H2AC12, H2AC19, H2AJ, H2AC14, H2AC20, H2AC17, H2AC7 |
| Response to metal ions | 0.004 | CSRP1 | Metalloprotease DUBs | 3.58e-08 | H2AC12, H2AC19, H2AC14, H2AC20, H2AC17, H2AC7 |
| Mitochondrial translation termination | 0.007 | MRPL4, MRPL47, MRRF | Packaging of Telomere Ends | 1.46e-06 | H2AC12, H2AC19, H2AJ, H2AC14, H2AC20, H2AC7 |
| NFE2L2 regulating antioxidant/detoxification enzymes | 0.009 | GCLC | HDACs deacetylate histones | 1.81e-06 | H2AC12, H2AC19, H2AC14, H2AC20, H2AC17, H2AC7 |
| RNA Polymerase I Transcription Initiation | 0.019 | EHMT2, POLR1D | DNA methylation | 1.95e-06 | H2AC12, H2AC19, H2AJ, H2AC14, H2AC20, H2AC7 |
| Synthesis of very long-chain fatty acyl-CoAs | 0.019 | ACSL3, HACD3 | RHO GTPase Effectors | 2.43e-06 | PIK3R4, S100A8, PRKACA, S100A9, RHOC, TUBB4A, H2AC12, H2AC19, H2AJ, H2AC14, H2AC20, H2AC7 |
| Membrane Trafficking | 0.021 | ACBD3, CALM, SCOC, AP4S1, MCFD2, ARF5, RABGAP1 | Recognition & association of DNA glycosylase with site containing an affected purine | 2.90e-06 | H2AC12, H2AC19, H2AJ, H2AC14, H2AC20, H2AC7 |
| KEAP1-NFE2L2 pathway | 0.023 | GCLC, PSME1 | Regulation of MECP2 expression and activity | 3.29e-06 | MECP2 |
| Loss of phosphorylation of MECP2 at T308 | 0.029 | CALM1 | Assembly of the ORC complex at the origin of replication | 3.29e-06 | H2AC12, H2AC19, H2AJ, H2AC14, H2AC20, H2AC7 |
| RUNX1 regulates transcription of genes involved in BCR signaling | 0.029 | ELF2 | HATs acetylate histones | 3.43e-06 | H2AC12, H2AC19, H2AJ, H2AC14, H2AC20, H2AC17, H2AC7, TAF9 |

## Discussion

The intricate molecular landscape of PDAC contributes to treatment inefficacy and a considerable rise in patient mortality rates (21). Inadequate early diagnostic and prognostic markers along with the ineffective current therapeutic options add to the factors that result in poor outcomes. There are still challenges in increasing the efficacy of immunotherapeutic strategies in PDAC. Identifying key targets that could regulate the immune response and their associated underlying mechanisms could help circumvent these (22). In this study, we found C3 and C5 to be upregulated in early PDAC but reduced in late-stage disease. We further demonstrated that their pharmacological inhibition increased cell proliferation and migration *in vitro*.

This study identified increased levels of complement *C3* and *C5* in early-stage (resectable) PDAC patients. PDAC cells hijack normal cellular processes to foster tumour growth and proliferation. One possible mechanism is by increasing the levels of C3 and C5 leading to the production of C3a and C5a and the subsequent recruitment of immune cells to the site of the tumour by chemotaxis (23). This immune response can then aid tumour growth by secreting cytokines like TNF-α and IL-6 which promote angiogenesis and subsequently supplies the growing tumour with nutrients and oxygen required for survival(24). Furthermore, the production of C3a and C5a, can attract immunosuppressors such as TGF-β and VEGF and suppressive myeloid cells to the tumour microenvironment and inhibit the anti-tumour functions of CD4 and CD8 cells (25). This implies that PDAC may increase C3 and C5 to support tumour growth and facilitate the development of a suppressive tumour microenvironment (TME).

A positive correlation between CRP and Complement C5 was observed. One study demonstrated that elevated levels of C5 can induce the production of pro-inflammatory cytokines and chemokines including TNF-α, IL-1β, and IL-6 (26), which are elevated in PDAC (27). These cytokines amplify the inflammatory response and activate signaling pathways, such as NF-κB and STAT3, that promote cell proliferation, survival and angiogenesis (4). Furthermore, CRP binds C5 to activate complement and secrete pro-inflammatory cytokines (28). The interaction between CRP and C5 may result in ROS species production which may induce DNA damage through dysregulated mitochondrial protein as shown in this study when complement is inhibited. Taken together, C5 and CRP may synergistically collaborate to fuel inflammatory processes that support tumour growth in PDAC.

Interestingly, C3 and C5 plasma levels were observed to be reduced in late-stage PDAC compared to early-stage PDAC. This may suggest an active immune response and activation of the complement pathway in the early stages of cancer (29). On the other hand, as PDAC progresses the immune system may become compromised leading to a decrease in C3 and C5 levels. We showed that inhibiting the complement pathway increases MIA PaCa-2 cells at the G0 phase and induces cell cycle arrest at the S phase by modulating the cell cycle-regulating protein Cyclin D1 which allowed the G1 to S phase transition seen in the increase of cells in the S phase following treatment, representing cancer progression. Studies have shown that, due to their genetic modification to proliferate faster, cancer cells minimise the time in the G0 phase of cell replication (30). Within the TME, complement proteins can activate key signaling pathways such as the PI3K/AKT, MAPK/ERK, or NF-κB pathways, involved in diverse cellular processes that promote tumorigenesis (31). Multiple studies have shown that inhibiting the complement pathway in ovarian cancer, colon cancer, and melanoma reduces tumour growth (7,32,33). However, the role of the complement pathway is not fully understood in PDAC. Studies have also suggested that decreased levels of C3 or C5 can disrupt these pathways, leading to changes in the cellular response and potentially increasing migration rates. Our findings together with other published studies suggest that the complement pathway may play context-dependent roles depending on the cancer type and potentially, tumour stage (8).

Proteomic analysis of the inhibition of the complement pathway demonstrated an increased expression of mitochondrial ribosomal proteins (MRPL4, MRPL47,) and mitochondrial ribosome recycling factor (MRRF). These proteins play crucial roles in protein synthesis, energy metabolism, respiration, and ATP synthesis. Most cancers alter mitochondrial functions to stimulate the Warburg effect, causing an increase in glucose production and uptake, fueling cancer growth, and an elevated ROS which could result in DNA damage and is indicative of inflammation (34). Moreover, pharmacological inhibition of complement downregulates H2A Clustered histone molecules which are responsible for connecting chromatin and regulating gene expression. Studies have shown that the dysregulation of these molecules may result in aberrant cell cycle progression, genome instability, DNA damage response as well as transcriptional regulation (35). Some studies suggest that pancreatic cancer cells may downregulate or decrease the expression of certain chromatin-remodelling proteins that are critical for DNA repair (36).

## Conclusion

The findings of this study demonstrated that early PDAC exhibits high expression levels of C3 and C5, which decrease as the severity of the tumour increases. Inhibition of the complement pathway was shown *in vitro* to result in a more aggressive phenotype by stimulating cellular growth, proliferation, and migration, indicating the involvement of complement C3 and C5 in tumour progression. Our findings suggest the dysregulation of the complement pathway in PDAC progression may further help in understanding the context-specific role of the complement pathway in carcinogenesis.

## Ethics clearance

Ethical clearance for this study was obtained from the Human Research Ethics Committee (Medical) of the University of Witwatersrand (M190734 and M140317). Before sample collection, all participating individuals gave written informed consent.

## Data Availability

All data produced in the present study are available upon reasonable request to the authors

## Acknowledgement

The authors acknowledge the clinical staff of the Hepatopancreatobiliary Unit at Chris Hani Baragwanath Hospital, Johannesburg, South Africa. Mrs Nnenna Elebo for assistance with plasma samples.

## Authors’ contributions

E.E.N conceptualised the study. Z.N, P.N, G.C, and E.E.N acquired funding for the project. Z.N, P.N, and E.E.N collected data. Z.N, P.N, J.D, J.OJ, M.S, T.N.A, G.C, and E.E.N performed data analysis and interpretation. Z.N, P.N, T.N.A and E.E.N wrote the initial draft. All authors critically reviewed and approved the final manuscript.

## Funding

The study was funded by the National Research Foundation grant (Grant number: 138367), the Cancer Association of South Africa (CANSA), and the University of the Witwatersrand Faculty Research Committee Individual Grant.

## Conflicts of Interest

The authors declare no conflict of interest.

---

**Figure S1:**
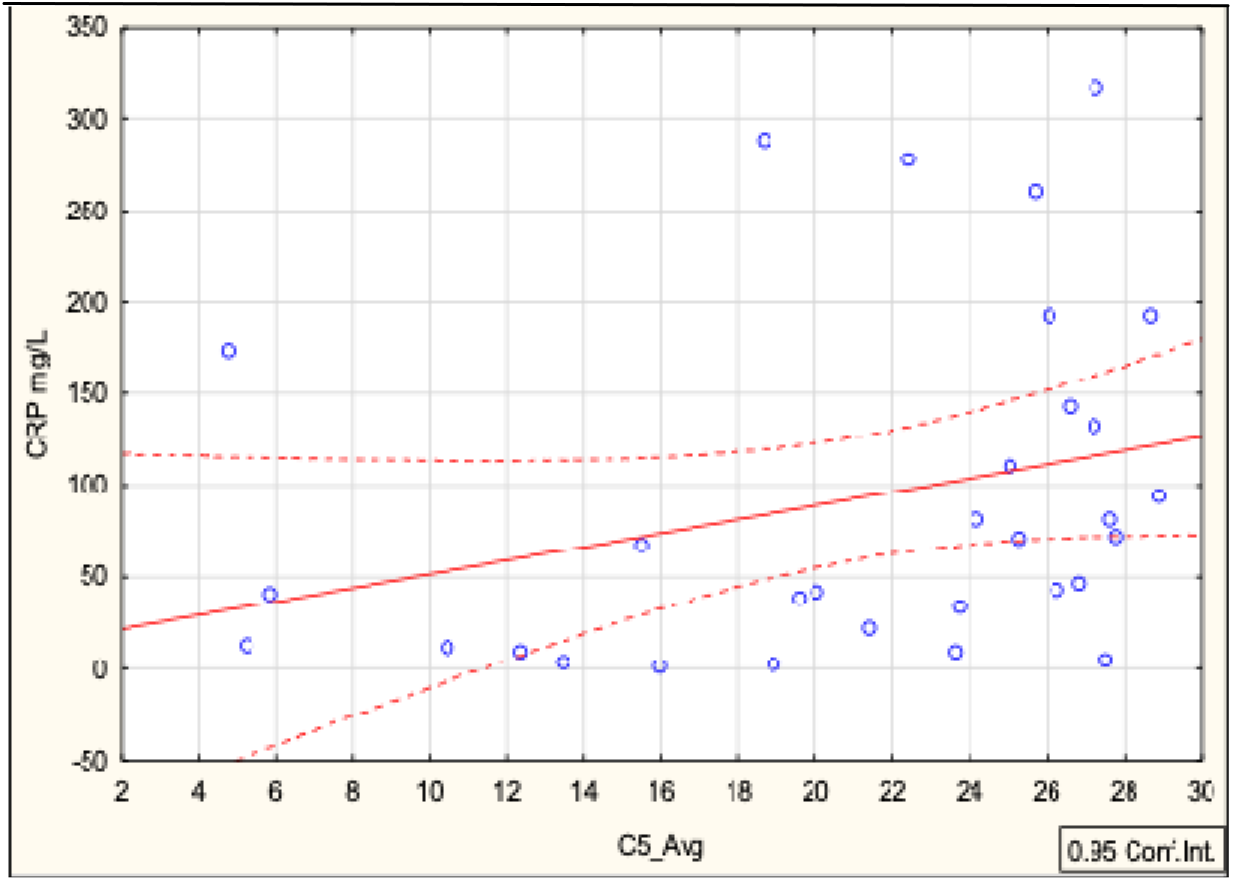
Complement C5 correlates with CRP. Scatter plot showing a positive correlation (r=0.45669) between C-reactive protein and the complement C5 which are both inflammatory markers (p=0.0098).

**Figure S2:**
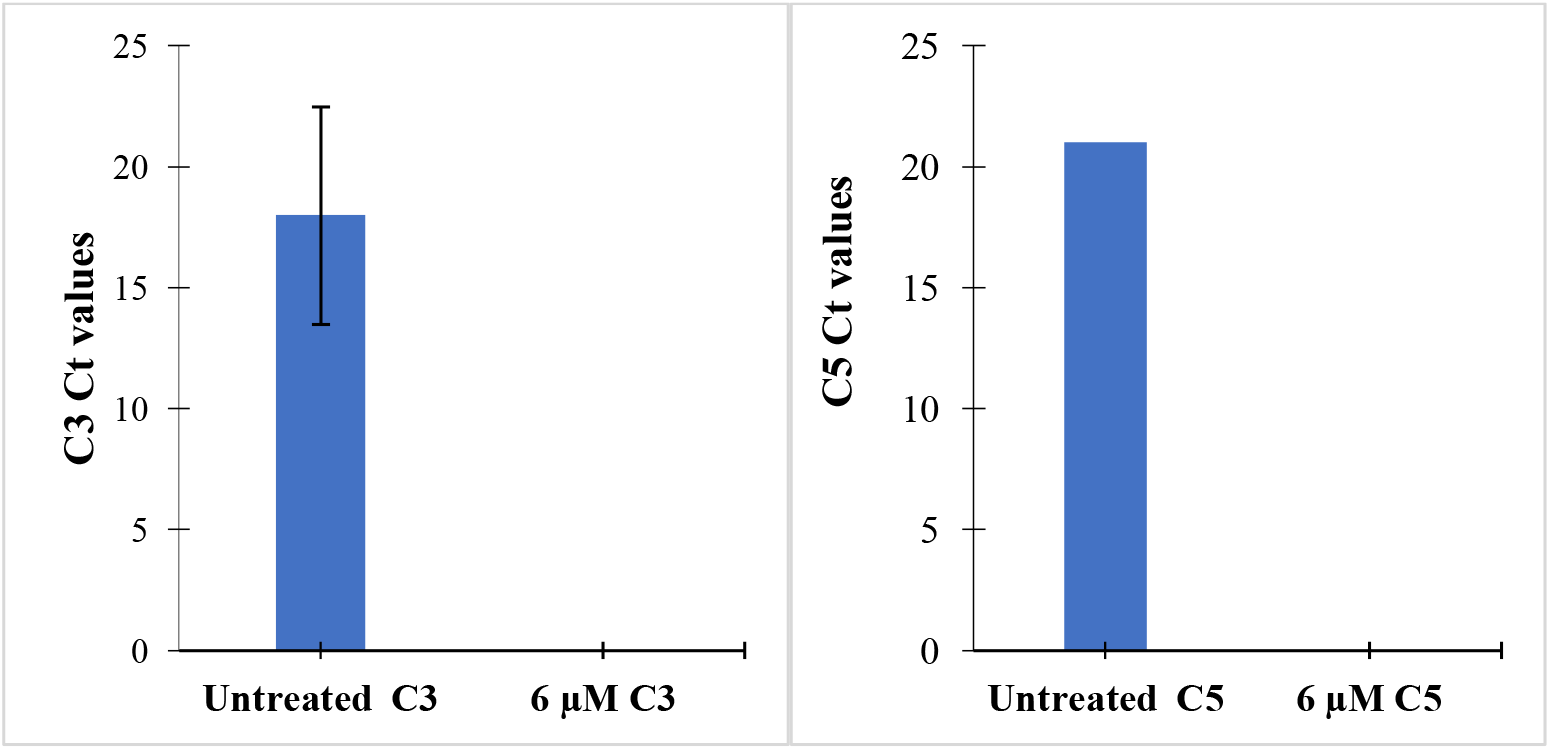
Confirmation of complement C3 and C5 inhibition using qPCR. Complement *C3* and *C5* are over-expressed in untreated MIA PaCa-2 cells and not expressed following treatment with compstatin.

**Table S1:** Demographic information for chronic pancreatitis and PDAC patients.

| Parameters | Chronic Pancreatitis<br>(n=6) | Resectable<br>(n=13) | Locally Advanced<br>(n=8) | Metastatic<br>(n=4) | p-value |
| --- | --- | --- | --- | --- | --- |
| Age, Median<br>[Range] | 51 [46 57] | 65 [54 66] | 56.5 [43 61] | 56 [46 76] |  |
| Gender |  |  |  |  | 0.367 |
| Male, n (%) | 6 (100) | 10 (77) | 6 (75) | 2 (50) |  |
| Female n (%) | 0 (0) | 3 (23) | 2 (25) | 2 (50) |  |
| Smoker |  |  |  |  | 0.685 |
| Yes n (%) | 5 (83.3) | 7 (53.2) | 4 (50) | 2 (50) |  |
| No n (%) | 1 (16.7) | 6 (46.2) | 4 (50) | 2 (50) |  |
| HIV Status |  |  |  |  | 0.650 |
| Positive n (%) | 1 (16.7) | 1 (7.7)) | 2 (25) | 0 (0) |  |
| Negative n (%) | 5 (83.3) | 12 (92.3) | 6 (75) | 4 (100) |  |
| Alcohol |  |  |  |  | 0.957 |
| Yes n (%) | 3 (50) | 5 (38.5) | 4 (50) | 2 (50) |  |
| No n (%) | 3 (50) | 8 (61.5) | 4 (50) | 2 (50) |  |
| Employment Status |  |  |  |  | 0.197 |
| Employed n (%) | 2 (33.3) | 1 (7.7) | 3 (37.5) | 0 |  |
| Unemployed n (%) | 3 (50) | 6 (46.15) | 4 (50) | 3 (75) |  |
| Retired n (%) | 0 (0) | 6 (46.15) | 1 (12.5) | 1 (25) |  |

**Table S2:**
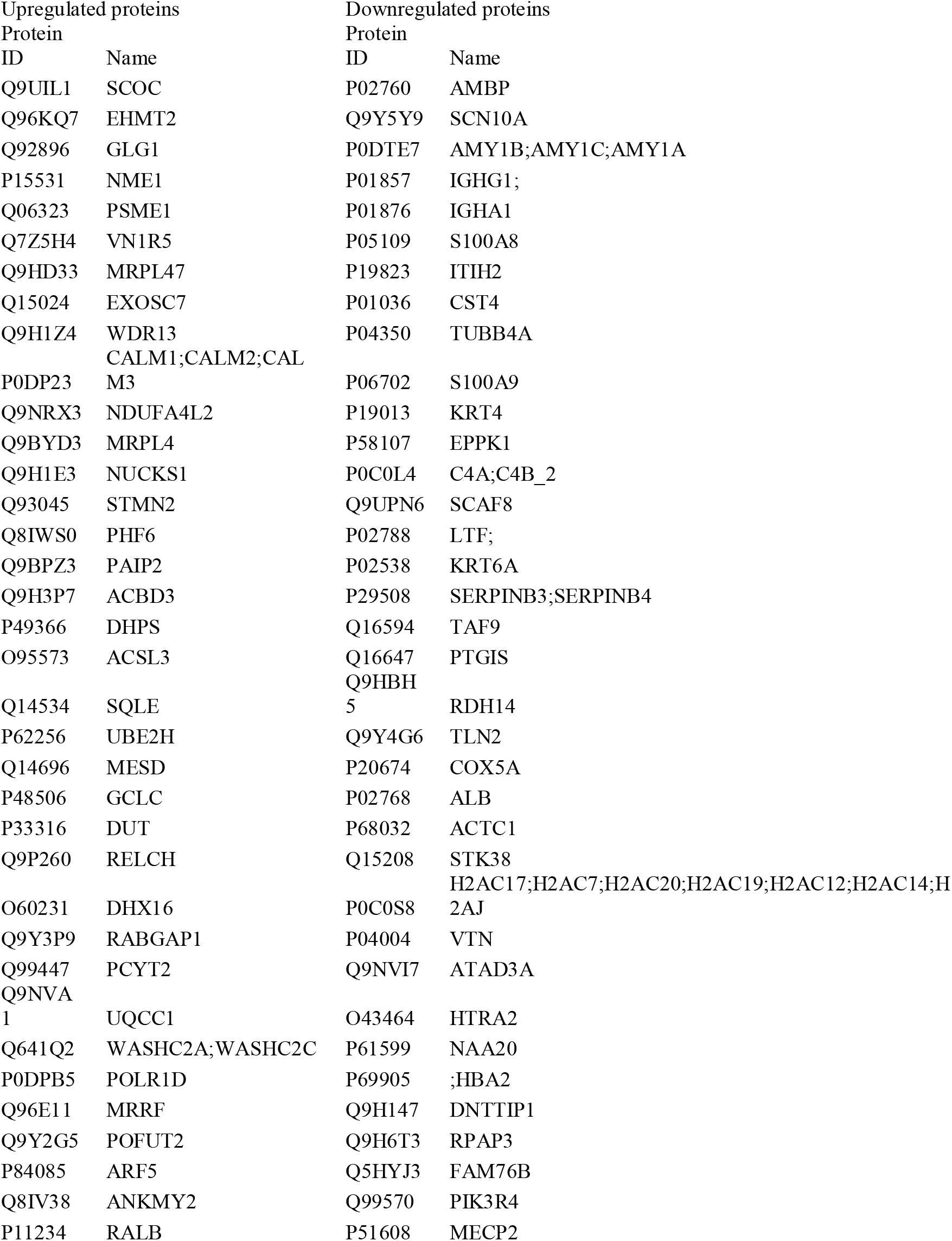
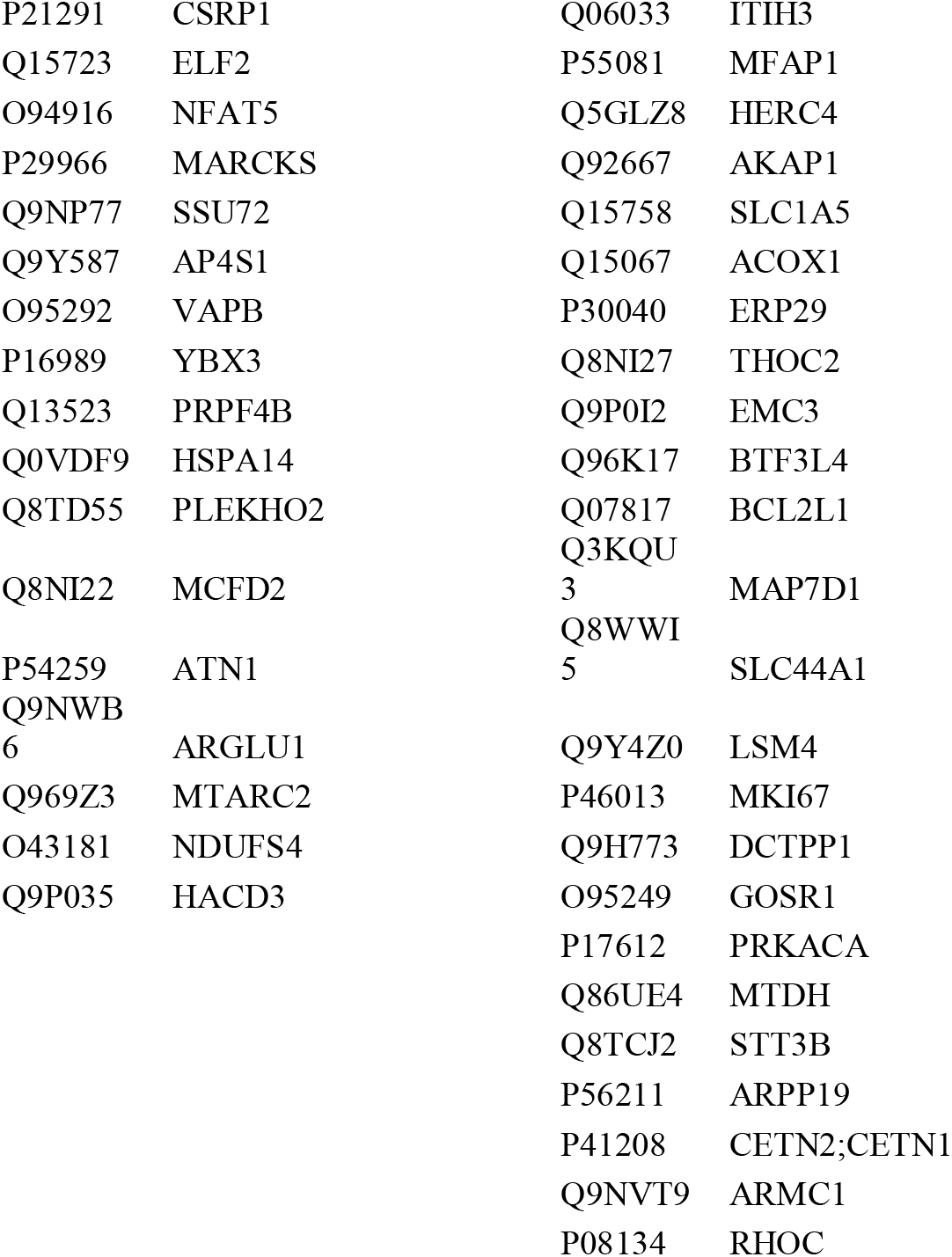
The list of up- and down-regulated proteins.

## Notes

### Competing Interest Statement

The authors have declared no competing interest.

### Author Declarations

Ethical clearance for this study was obtained from the Human Research Ethics Committee (Medical) of the University of Witwatersrand (M190734 and M140317).

